# Harnessing Transcriptomic Signals for Amyotrophic Lateral Sclerosis to Identify Novel Drugs and Enhance Risk Prediction

**DOI:** 10.1101/2023.01.18.23284589

**Authors:** Oliver Pain, Ashley Jones, Ahmad Al Khleifat, Devika Agarwal, Dzmitry Hramyka, Hajer Karoui, Jędrzej Kubica, David J. Llewellyn, Janice M. Ranson, Zhi Yao, Alfredo Iacoangeli, Ammar Al-Chalabi

## Abstract

**Introduction:** Amyotrophic lateral sclerosis (ALS) is a fatal neurodegenerative disease. This study integrates the latest ALS genome-wide association study (GWAS) summary statistics with functional genomic annotations with the aim of providing mechanistic insights into ALS risk loci, inferring drug repurposing opportunities, and enhancing prediction of ALS risk and clinical characteristics.

**Methods:** Genes associated with ALS were identified using GWAS summary statistic methodology including SuSiE SNP-based fine-mapping, and transcriptome- and proteome-wide association study (TWAS/PWAS) analyses. Using several approaches, gene associations were integrated with the DrugTargetor drug-gene interaction database to identify drugs that could be repurposed for the treatment of ALS. Furthermore, ALS gene associations from TWAS were combined with observed blood expression in two external ALS case-control datasets to calculate polytranscriptomic scores and evaluate their utility for prediction of ALS risk and clinical characteristics, including site of onset, age at onset, and survival.

**Results:** SNP-based fine-mapping, TWAS and PWAS identified 117 genes associated with ALS, with TWAS and PWAS providing novel mechanistic insights. Drug repurposing analyses identified five drugs significantly enriched for interactions with ALS associated genes, with directional analyses highlighting α-glucosidase inhibitors may exacerbate ALS pathology. Additionally, drug class enrichment analysis showed calcium channel blockers may reduce ALS risk. Across the two observed expression target samples, ALS polytranscriptomic scores significantly predicted ALS risk (*R*^2^ = 4%; *p*-value = 2.1×10^−21^).

**Conclusions:** Functionally-informed analyses of ALS GWAS summary statistics identified novel mechanistic insights into ALS aetiology, highlighted several therapeutic research avenues, and enabled statistically significant prediction of ALS risk.

## Introduction

Amyotrophic lateral sclerosis (ALS), also known as motor neurone disease, is a neurodegenerative disease affecting both upper and lower motor neurones, leading to progressive loss of voluntary muscle control, and ultimately respiratory failure within 3-5 years after disease onset (Van Es et al., 2017). The lifetime risk of ALS is 1 in 300 and a mean age of onset of 56 years, making it the most common neurodegenerative midlife disease (Brown & Al-Chalabi, 2017; Johnston et al., 2006). There is no diagnostic test for ALS, and substantial heterogeneity in clinical presentation and speed of progression make diagnosis of ALS challenging (Bendotti et al., 2020). ALS is currently diagnosed based on a series of tests ruling out other diseases, alongside a detailed history of symptoms observed by a physician, and a full medical history. The U.S. Food and Drug Administration has currently approved three drugs for the treatment of ALS, including Riluzole, Edavarone and Relyvrio. However, these drugs have a limited effect on life expectancy and daily functioning (Cruz, 2018; Heo, 2022; Miller et al., 2012). Further development of pharmacological treatments for ALS is therefore needed.

There is substantial evidence that genetic factors play an important role in the aetiology of ALS. Between 5-20% of people with ALS have a clear family history, referred to as familial ALS, with non-familial cases of ALS referred to as sporadic ALS (Al-Chalabi et al., 2017). Within familial cases of ALS, several rare genetic variants with large effect on ALS risk have been identified, such as within genes *C9orf72* (DeJesus-Hernandez et al., 2011) and *TBK1* (Freischmidt et al., 2015). Sporadic ALS cases also have a strong genetic aetiology, with twin-based heritability estimates between 40-60%, and ∼20% of those with the sporadic disease carrying known ALS risk variants (Al-Chalabi et al., 2010; McLaughlin et al., 2015; Mehta et al., 2022). In addition to the rare large effect variants associated with ALS, there is evidence of a substantial common genetic component to ALS risk, with SNP-based heritability estimates of 21% (Keller et al., 2014). The largest genome-wide association study (GWAS) of ALS (29,612 people with ALS and 122,656 controls) recently reported 15 genome-wide significant loci associated with ALS risk (van Rheenen et al., 2021), replicating previous associations and identifying novel risk loci. The authors analysed each significant locus to highlight likely causal genetic variation using statistical fine-mapping and inferred the underlying molecular mechanisms through the integration of functional genomic annotations such as expression quantitative trait loci (eQTL) data.

Transcriptome-wide association study (TWAS), a powerful approach for the integration of eQTL data with GWAS summary statistics, infers up- and down-regulation of gene expression associated with the GWAS phenotype (Gusev et al., 2016). This approach provides valuable insights into the molecular mechanisms underlying genome-wide significant loci, and due to the aggregation of associations across variants and a reduced multiple testing burden, TWAS can identify significant associations within loci not previously achieving genome-wide significance in the GWAS (Dall’Aglio et al., 2020; Gusev et al., 2018). A key advantage of TWAS over traditional studies analysing observed differential gene expression in cases and controls is the gain in power by combining the large sample sizes of GWAS with the precious and smaller eQTL datasets. This is particularly relevant for brain-related disorders for which the disease relevant tissue is only accessible post-mortem. The TWAS framework has recently been expanded to other molecular traits, such as altered protein-level data (pQTL), referred to as proteome-wide associations study (PWAS), uncovering unique insights into the downstream effects of disease associated loci (Wingo et al., 2021).

TWAS and PWAS identify genes associated with the GWAS phenotype, but also provide the direction of effect on the phenotype, as well as some degree of spatiotemporal specificity depending on the samples used to derive TWAS/PWAS models (Huckins et al., 2019). This contrasts with functionally agnostic gene association analyses, such as MAGMA (de Leeuw et al., 2015), which aggregate genetic associations within gene regions, thereby providing no information on direction of effect or spatiotemporal context. The additional information available from TWAS/PWAS can be used to enhance downstream analyses, such as drug repurposing analysis and prediction modelling.

Drug enrichment analysis using unsigned gene association summary statistics is a commonly used approach to identify drugs for repurposing (Gaspar et al., 2018). However, no information regarding direction of effect is considered, so it is possible this approach might identify drugs that exacerbate pathological mechanisms underlying the GWAS phenotype. In contrast, TWAS/PWAS results can be used to provide insight into whether a given drug interacts with disease associated genes that reduce disease risk specifically (So et al., 2017).

Another key application of GWAS is phenotype prediction, primarily using polygenic scores, calculated as the GWAS-effect size weights sum of associated alleles (Choi et al., 2020). A limitation of polygenic scores is their limited predictive utility, typically only capturing a fraction of the SNP-based heritability. TWAS/PWAS results provide an alternative approach for phenotype prediction, as they can be used in combination with observed expression/protein data in a target sample to calculate omic-based scores predicting the GWAS phenotype (Marigorta et al., 2017). When using TWAS results in combination with observed expression in a target sample, the resulting scores can be referred to as polytranscriptomic scores (PTS). PTS leverage the power of GWAS and capture both genetic and environmental risk an individual may carry, broadening the phenotypic variance that can be explained over polygenic scores. We sought to further utilise the GWAS summary statistics from the latest ALS GWAS, along with the latest functional genomic annotations. Here, we perform TWAS and PWAS of ALS to identify novel genetic associations with ALS, identify drug repurposing opportunities for ALS, and evaluate the predictive utility of PTS for ALS risk and clinical characteristics.

## Methods

An overview of the study design is shown in Figure 1. Using the latest ALS GWAS summary statistics, we performed a series of analyses to identify genes associated with ALS. Gene-finding analyses include SNP-based fine-mapping, integration of expression and protein data (TWAS and PWAS), and gene association analysis using MAGMA. The results of these analyses were used for three downstream aims. First, identification of high-confidence gene associations with ALS was achieved using results of SNP-based fine-mapping, TWAS and PWAS. Second, identification of novel drug repurposing opportunities for ALS was based on results from TWAS and MAGMA. Third, the results of TWAS were used to calculate PTS, which we then evaluated in two external blood expression ALS case-control datasets. More information regarding these analyses is provided below.

**Figure 1.**
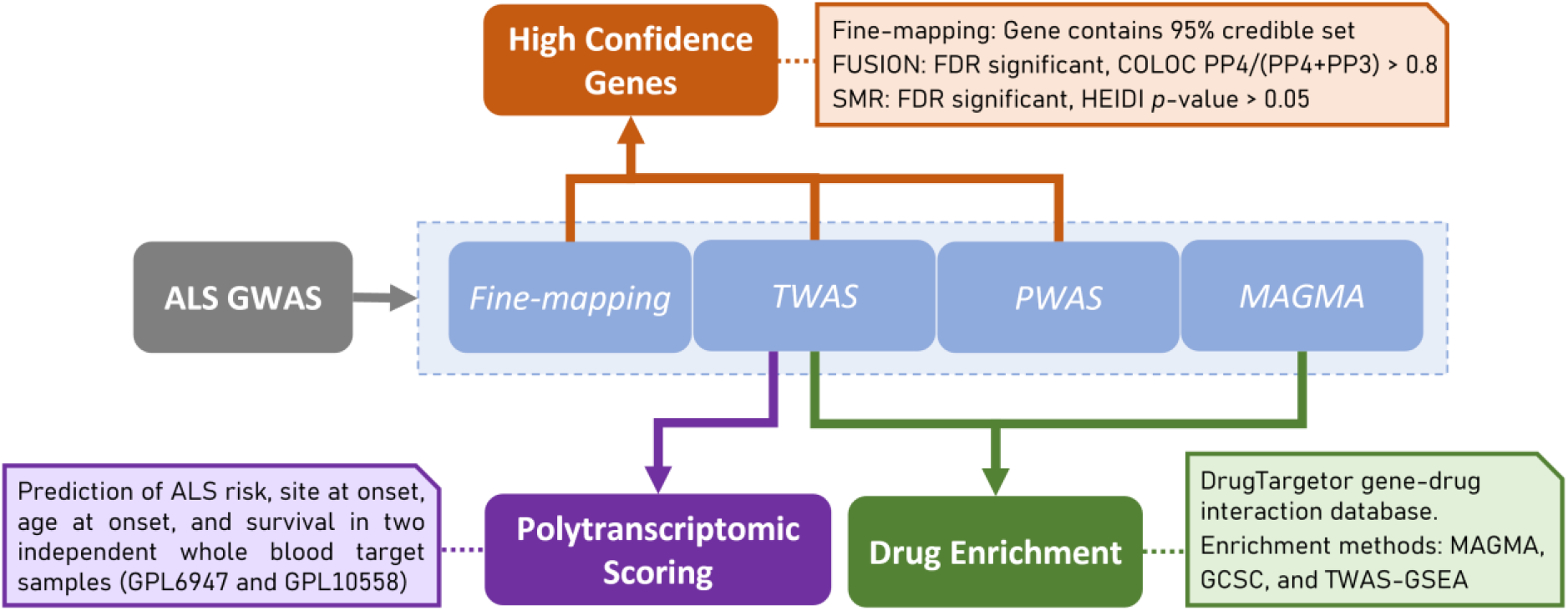
Schematic representation of study design and statistical analysis.

### ALS GWAS summary statistics

We used the most recent ALS GWAS summary statistics, which are publicly available (van Rheenen et al., 2021). The European ancestry-only GWAS summary statistics were used to avoid linkage disequilibrium (LD) mismatch with external reference data downstream, all of which is derived within European populations. The European-only ALS GWAS included 27,205 people with ALS and 110,881 control participants. GWAS summary statistics underwent standard quality control (see Supplementary Information 1).

### Gene finding analyses

#### Fine-mapping

The SuSiE R package was used to perform variant-level statistical fine-mapping of ALS GWAS summary statistics (Wang et al., 2020). SuSiE calculates the posterior inclusion probability of genetic variants being the casual variant for each genome-wide significant locus. SuSiE was performed assuming a single causal variant (L = 1) to avoid LD mismatch issues between the GWAS sample and 1KG European reference population. As a sensitivity analysis, SuSiE was also performed allowing up to 10 independent causal signals. Further information is available in the Supplementary Information 2.

#### TWAS/PWAS

##### Expression and protein panels

Given ALS-associated genes are primarily expressed in brain tissues (van Rheenen et al., 2021), we inferred differential expression/protein levels associated with ALS using a range of brain expression/protein panels (Table 1). Furthermore, we included blood expression panels, as these often have a larger sample size available than brain tissues, and blood eQTL data can be used as a proxy for brain tissues due to the moderate correlation between eQTL effects across tissues (GTEx Consortium, 2015). TWAS and PWAS were performed using FUSION software (https://github.com/gusevlab/fusion_twas). Most panels were previously derived and are publicly available via the FUSION website, though we generated novel TWAS models from post-mortem primary motor cortex RNA-seq and genotype data from the King’s College London (KCL) and Medical Research Council London Neurodegenerative Diseases Brain Bank, herein referred to as ‘KCL Brain Bank’ (Iacoangeli et al., 2021; Jones et al., 2021). KCL Brain Bank contains a mixture of people with ALS and controls, and FUSION software was used to generate the TWAS models after stringent quality control (see Supplementary Information 3).

**Table 1.**
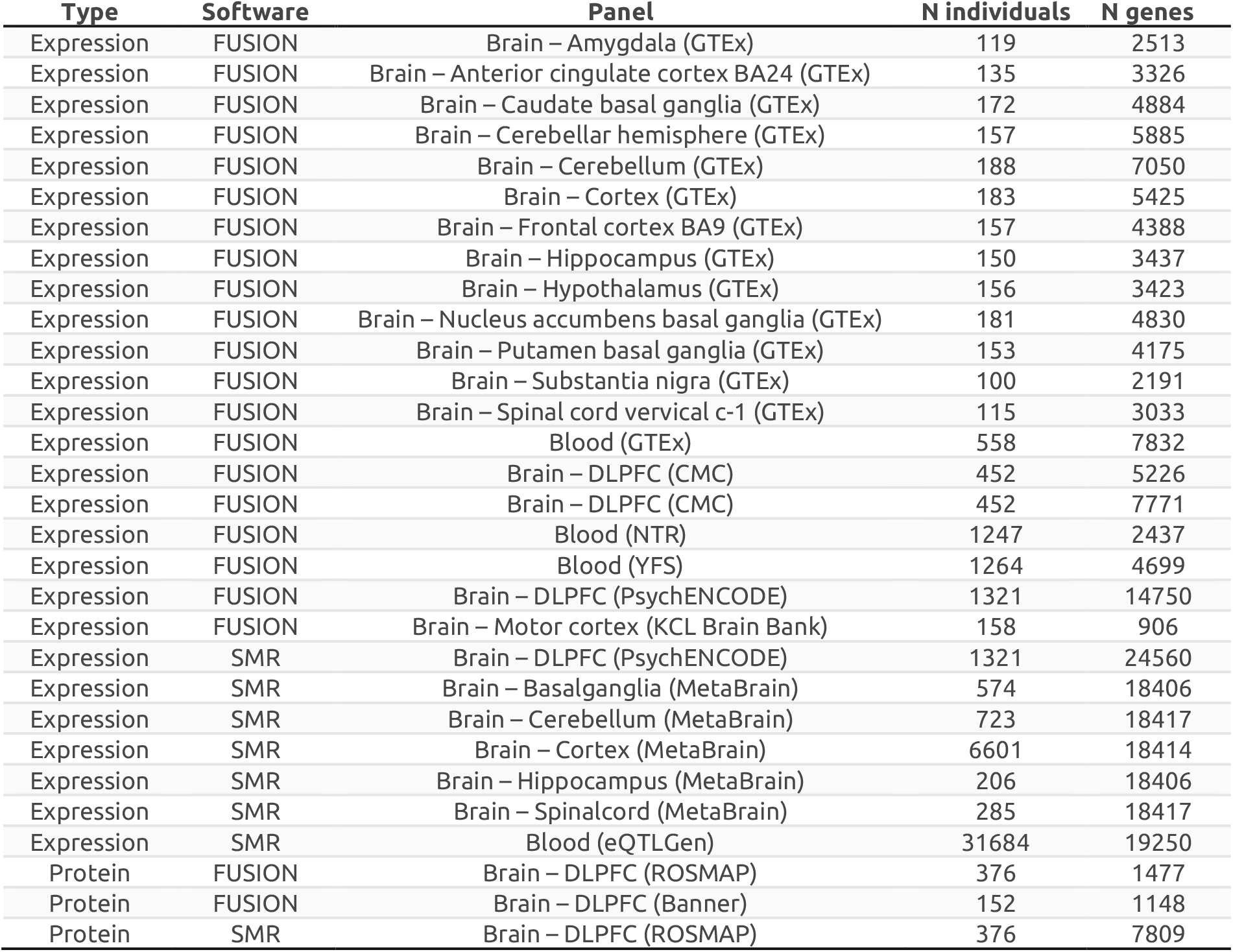
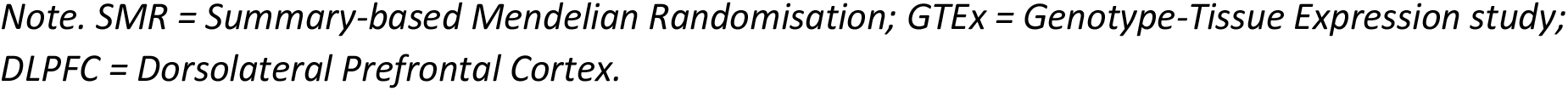
Expression and protein panels used for TWAS and PWAS.

| Type | Software | Panel | N individuals | N genes |
| --- | --- | --- | --- | --- |
| Expression | FUSION | Brain – Amygdala (GTEx) | 119 | 2513 |
| Expression | FUSION | Brain – Anterior cingulate cortex BA24 (GTEx) | 135 | 3326 |
| Expression | FUSION | Brain – Caudate basal ganglia (GTEx) | 172 | 4884 |
| Expression | FUSION | Brain – Cerebellar hemisphere (GTEx) | 157 | 5885 |
| Expression | FUSION | Brain – Cerebellum (GTEx) | 188 | 7050 |
| Expression | FUSION | Brain – Cortex (GTEx) | 183 | 5425 |
| Expression | FUSION | Brain – Frontal cortex BA9 (GTEx) | 157 | 4388 |
| Expression | FUSION | Brain – Hippocampus (GTEx) | 150 | 3437 |
| Expression | FUSION | Brain – Hypothalamus (GTEx) | 156 | 3423 |
| Expression | FUSION | Brain – Nucleus accumbens basal ganglia (GTEx) | 181 | 4830 |
| Expression | FUSION | Brain – Putamen basal ganglia (GTEx) | 153 | 4175 |
| Expression | FUSION | Brain – Substantia nigra (GTEx) | 100 | 2191 |
| Expression | FUSION | Brain – Spinal cord cervical c-1 (GTEx) | 115 | 3033 |
| Expression | FUSION | Blood (GTEx) | 558 | 7832 |
| Expression | FUSION | Brain – DLPFC (CMC) | 452 | 5226 |
| Expression | FUSION | Brain – DLPFC (CMC) | 452 | 7771 |
| Expression | FUSION | Blood (NTR) | 1247 | 2437 |
| Expression | FUSION | Blood (YFS) | 1264 | 4699 |
| Expression | FUSION | Brain – DLPFC (PsychENCODE) | 1321 | 14750 |
| Expression | FUSION | Brain – Motor cortex (KCL Brain Bank) | 158 | 906 |
| Expression | SMR | Brain – DLPFC (PsychENCODE) | 1321 | 24560 |
| Expression | SMR | Brain – Basalganglia (MetaBrain) | 574 | 18406 |
| Expression | SMR | Brain – Cerebellum (MetaBrain) | 723 | 18417 |
| Expression | SMR | Brain – Cortex (MetaBrain) | 6601 | 18414 |
| Expression | SMR | Brain – Hippocampus (MetaBrain) | 206 | 18406 |
| Expression | SMR | Brain – Spinalcord (MetaBrain) | 285 | 18417 |
| Expression | SMR | Blood (eQTLGen) | 31684 | 19250 |
| Protein | FUSION | Brain – DLPFC (ROSMAP) | 376 | 1477 |
| Protein | FUSION | Brain – DLPFC (Banner) | 152 | 1148 |
| Protein | SMR | Brain – DLPFC (ROSMAP) | 376 | 7809 |

*Note. SMR = Summary-based Mendelian Randomisation; GTEx = Genotype-Tissue Expression study; DLPFC = Dorsolateral Prefrontal Cortex.*

##### FUSION/SMR

We performed TWAS and PWAS using both FUSION and summary-based mendelian randomisation (SMR) methods to utilise the resources available for each of these methods and compare results across methods (see Supplementary Information 4). Both SMR and FUSION test whether genetic variants associated with the GWAS phenotype are also associated with differential gene expression or protein levels, using a subsequent colocalisation analysis to test whether the overlapping association is driven by the same causal variant. Further details can be found in Supplementary Information 4. False discovery rate (FDR) correction for multiple testing was performed across all expression panels for FUSION and SMR separately.

#### MAGMA gene association analysis

MAGMA v1.10 was also used to calculate gene associations (de Leeuw et al., 2015). MAGMA estimates gene associations by calculating the mean association of variants within gene regions, accounting for LD using Brown’s method. We defined gene locations using the NCBI37.3 locations file on the MAGMA website (https://ctg.cncr.nl/software/magma), using a 35 kb upstream and 10 kb downstream gene window, to allow for regulatory regions around each gene. We used the 1KG Phase 3 European LD reference available on the MAGMA website.

### Defining high-confidence genes

We used the following criteria to define genes associated with ALS with ‘high confidence’: Gene contains all variants within SuSiE 95% credible set, FDR significant TWAS/PWAS association and colocalised (coloc software PP4/(PP3 + PP4) > 0.8), FDR significant SMR association and colocalised (HEIDI p-value > 0.05). The threshold used to define colocalisation by coloc was chosen to highlight the probability that the association is driven by the same causal variant (PP4) rather than due to linkage (PP3). TWAS will have already determined an association in the locus for both traits and we can ignore the posterior probability of models where there is no association for either or both traits. Significant MAGMA gene associations are not considered high confidence as results are liable to confounding due to LD.

### Drug repurposing analysis

#### Enrichment methods

We tested for enrichment of drug-gene interactions using three approaches: MAGMA, Gene Co-regulation Score Regression (GCSC), and TWAS-based Gene Set Enrichment Analysis (TWAS-GSEA).

MAGMA gene-set enrichment analysis is based on MAGMA estimated gene associations and binary drug-gene interaction data (de Leeuw et al., 2015). GCSC regression is a method that leverages gene co-regulation to test for and enrichment of TWAS associations within gene-sets or associated with gene-properties (Siewert-Rocks et al., 2022). TWAS-GSEA is based on TWAS estimated gene associations and directional drug-gene interaction data (Pain et al., 2019).

A key distinction between these methods is that MAGMA and GCSC do not account for the direction of association between the gene, the GWAS phenotype, and the drug. Therefore, when testing for an enrichment of genes that interact with a given drug, they fail to test whether the drug would decrease risk of ALS. In contrast, TWAS-GSEA does consider the direction of association between the gene, phenotype, and drug. As a result, enriched drugs using this method are suggested to induce expression changes reducing risk of ALS. Further details on these methods and their differences are provided in the Supplementary Information 5.

#### Drug-gene interaction data

DrugTargetor is a tool for identifying drugs that interact with genes associated with a GWAS phenotype (Gaspar et al., 2018). DrugTargetor uses a drug-gene interaction dataset drawn from a range of sources including ChEMBL, PHAROS, PDSP Ki database and NCBI PubChem BioAssay (https://github.com/hagax8/drugtargetor/blob/master/wholedatabase_for_targetor). The DrugTargetor dataset indicates whether a drug interacts with a given gene’s expression or protein product, and whether the interaction leads to an increase or a decrease in activity. We used MAGMA, GCSC and TWAS-GSEA to test for enrichment of drugs based on the DrugTargetor drug-gene interaction dataset. Binary gene sets indicating drug-gene interactions were used for MAGMA and GCSC, as these methods do not account for the direction of effect. For TWAS-based enrichment, we coded drug-interactions as -1 or 1 to indicate whether the drug decreased (labelled as ‘DECREASED_EXPRESSION’, ‘NEGATIVE_RESPONSE’, or ‘OPPOSITE_RESPONSE’) or increased (labelled as ‘INCREASED_EXPRESSION’ or ‘POSITIVE_RESPONSE’) the activity of the gene. Otherwise, drug-gene interactions were coded as 0 where there was no directional evidence of drug-gene interaction. Drug enrichment analyses were restricted to drugs interacting with at least 2 genes with available association statistics.

#### Anatomical Therapeutic Chemical (ATC) enrichment analysis

After estimating enrichment of specific drugs, we tested whether drugs with specific level 3 ATC (pharmacological subgroup) codes were enriched. This was carried out using the non-parametric Wilcoxon test, testing whether drugs within a given ATC group were enriched for association compared to all other drugs available in DrugTargetor. A one-sided test was used for MAGMA and GCSC, but a two-sided test was used for TWAS-GSEA to test direction and significance of ATC group enrichments. FDR correction was used to account for multiple testing when determining statistical significance. ATC enrichment was restricted to ATC groups containing at least 5 drugs with drug enrichment statistics available.

### Polytranscriptomic scoring (PTS)

#### Target gene expression datasets

To assess the utility of PTS for predicting ALS risk and clinical features, we used two whole blood gene expression datasets from the Gene Expression Omnibus. These datasets are a part of the same Gene Expression Omnibus series (GSE112681) but separated due to the use of two array platforms to measure gene expression. The datasets corresponding to each platform are termed GPL6947 and GPL10558. These datasets both consist of people diagnosed with ALS and controls, and were originally used for a differential gene expression study of ALS (Van Rheenen et al., 2018). The expression and phenotype data were downloaded using GEOquery. The expression data was averaged across probes for each gene using the limma R package ‘avereps’ function (Ritchie et al., 2015), and values were then log2 transformed. The following phenotypes were available for both datasets: ALS case-control status, site of onset (spinal vs bulbar), age at onset (years), and survival.

#### Calculation of Polytranscriptomic Scores

An analogous approach to calculating polygenic scores can be used to calculate PTS. Polygenic scores are typically calculated as the sum of alleles weighted by their GWAS effect size. In contrast, PTS are calculated as the sum of observed expression values weighted by their TWAS effect size. Specifically, PTS were calculated as the sum of gene expression Z-scores, weighting each gene by the corresponding TWAS Z-score.

Analogous to the commonly used LD-based clumping and *p*-value thresholding for calculating polygenic scores, we accounted for the non-independence of nearby TWAS associations by clumping based on a predicted gene expression correlation matrix, and applied a range of *p*-value thresholds to select different numbers of genes to be included in the PTS. The predicted expression correlation matrix was generated by predicting expression into the European subset of the 1KG reference sample using the SNP-weights in the corresponding TWAS models, and then calculating the Pearson correlation between all genes within a 500kb window. We removed TWAS associations if they had a high Pearson correlation with a lead gene (*r* > 0.95). This clumping and thresholding approach was carried out using the IFRisk script (https://github.com/opain/Inferred-functional-risk-scoring) and is consistent with a previous study generating TWAS-based risk scores (Pain et al., 2021).

We generated PTS using TWAS results from all panels combined, and TWAS results from brain panels and blood panels separately. We also generated PTS using only TWAS associations that showed evidence of colocalisation (PP4 > 0.8). We only considered the colocalisation PP4 value to define colocalisation for this analysis as the PP4-PP3 ratio may be unstable for genes at lower TWAS p-value thresholds. We averaged TWAS Z-scores for a given gene if the gene was in multiple panels after clumping.

#### PTS association analysis

We compared ALS PTS between people with ALS and healthy controls (binary) and compared the ALS PTS across clinical characteristics within people diagnosed with ALS alone, including spinal versus bulbar site of onset (binary), age at onset (continuous) and survival (continuous). We used linear regression to test the PTS association for all outcomes, converting the observed *R*^2^ to the liability scale for binary outcomes. For case-control analysis, we assumed a population prevalence of 1/300. For spinal-bulbar onset analysis, when converting to the liability scale we assumed an arbitrary population prevalence of 0.5 to aid comparison with future studies. We included sex as a covariate throughout.

We performed regression within each gene expression platform (GPL6947 and GPL10558) separately, and then meta-analysed results using inverse variance weighting.

#### Observed differential expression analysis

In addition to TWAS, which infers differential expression associated with ALS risk, we also estimated the observed evidence of differential expression associated with ALS within the GPL6947 and GPL10558 ALS case-control cohorts. This was carried out to enable comparison of TWAS-inferred and observed differential expression, as this can highlight observed differential gene expression that is a consequence of the disease (Marigorta et al., 2017).

We tested the Pearson correlation between ALS case-control status and observed expression levels within the GPL6947 and GPL10558 cohorts separately, and subsequently meta-analysed using inverse-variance weighting. FDR correction was used to account for multiple testing when determining statistical significance.

## Results

### Gene discovery

We performed SNP fine-mapping, TWAS and PWAS based on ALS GWAS summary statistics to define a set of high-confidence genes associated with ALS.

SNP-based fine-mapping, with an assumption of a single causal signal, identified five high-confidence genes (details in Supplementary Information 6 and Tables S1-S2). TWAS using FUSION and SMR identified 108 genes with significant differential expression in ALS and strong evidence of colocalisation (Tables S3-S4). PWAS using FUSION and SMR identified altered levels of 7 proteins in ALS and strong evidence of colocalisation, three of which were also identified as high confidence using TWAS (Figure 2, Tables S5-S6). Across SNP fine-mapping, TWAS and PWAS analyses, 117 unique genes were identified as high-confidence associations. Further details of gene discovery results are in the Supplementary Information 6 and Figure S1.

**Figure 2.**
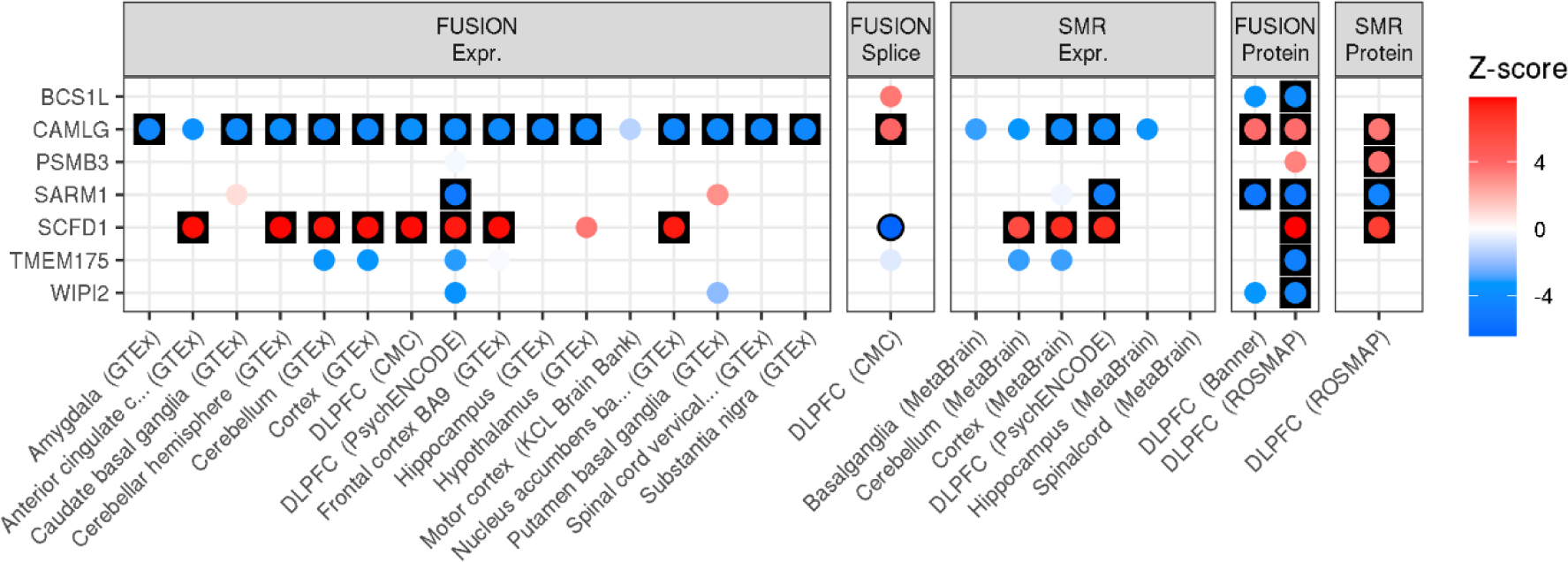
TWAS and PWAS associations for high-confidence ALS-associated genes defined using PWAS. Results are only shown for brain tissue expression and protein panels. Results are separated by the method and external data used. FUSION and SMR results are shown for all panels, with each point coloured according to the Z-score of association. Red indicates an increased expression/protein level in people diagnosed with ALS, and blue indicates decreased expression/protein level in people diagnosed with ALS. Results have a black outline if the association was FDR significant, and are in a black square if the association was FDR significant and showed evidence of colocalisation.

### Drug repurposing for ALS

We used MAGMA, GCSC and TWAS-GSEA to test for enrichment of ALS-associated genes that interact with specific drugs (Figure 3). GCSC identified five significantly enriched drugs, including acarbose (FDR = 1.03×10^−11^), voglibose (FDR = 2.09×10^−10^), sertindole (FDR = 5.99×10^−7^), oxybuprocaine (FDR = 8.31×10^−5^), and ethambutol (FDR = 2.95×10^−3^). MAGMA and TWAS-GSEA did not identify any significantly enriched drugs after multiple testing correction. Of the genes identified by GCSC, TWAS-GSEA found nominally significant evidence across multiple expression panels that acarbose and voglibose induce an expression signature that may increase risk of ALS. Although non-significant, TWAS-GSEA results for sertindole and ethambutol indicated these drugs induced an expression profile that was associated with a reduced risk of ALS.

**Figure 3.**
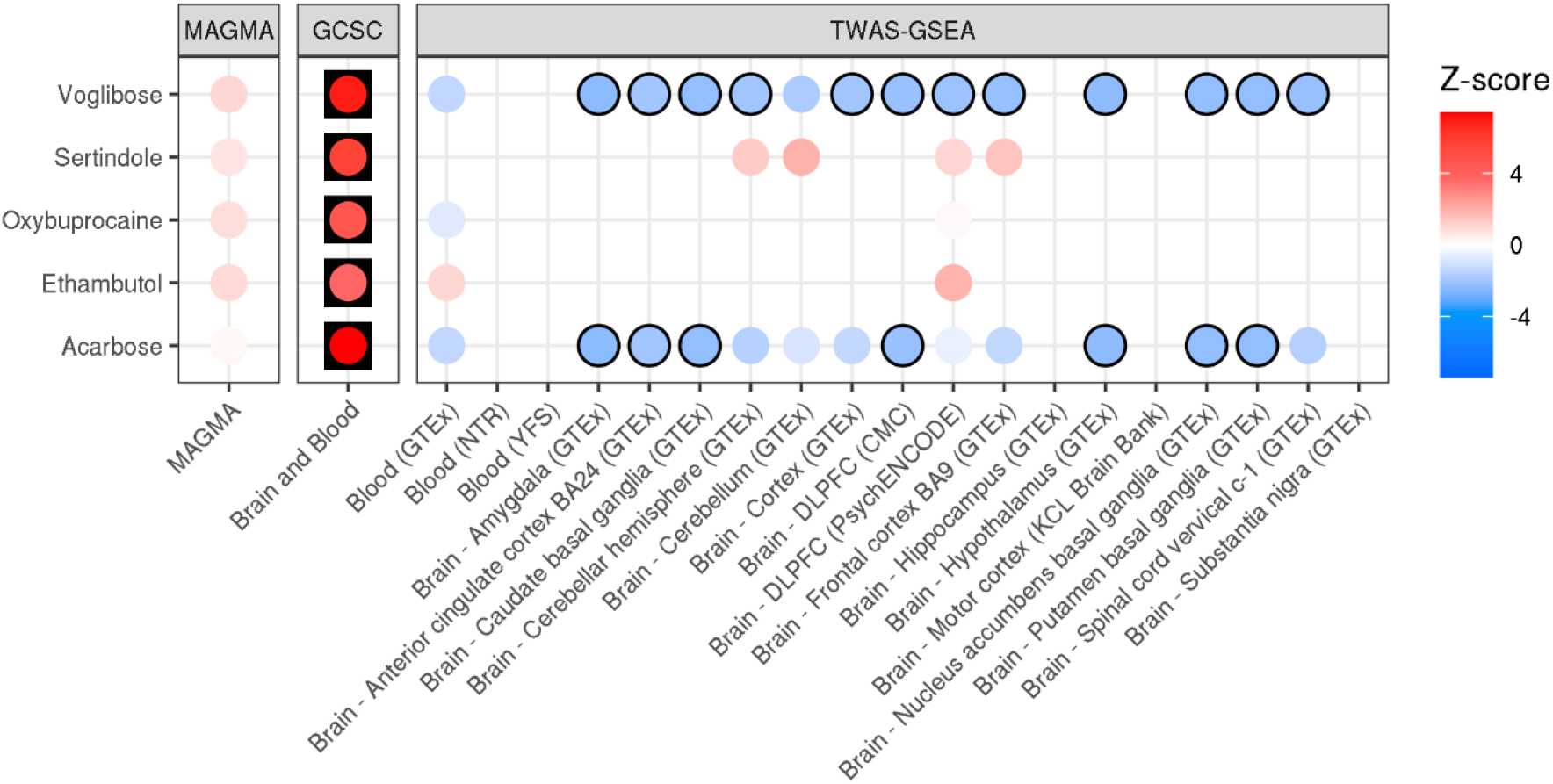
Drug enrichment results for ALS using MAGMA, GCSC and TWAS-GSEA. Each point is coloured according to the Z-score of enrichment. For TWAS-GSEA, positive Z-scores (red) indicates the drug reduces risk of ALS, whereas negative Z-scores (blue) indicate the drug increases risk of ALS. Results have a black outline if the association was nominally significant, and are in a black square if the association was FDR significant.

We then tested for enrichment of ATC classes based on the drug associations from MAGMA, GCSC and TWAS-GSEA (Figure 4). Using MAGMA results identified 2 significant ATC categories, including ‘antidepressants’ (FDR = 3.55×10^−6^) and ‘antipsychotics’ (FDR = 3.02×10^−3^). Using GCSC results, only the ATC group ‘selective calcium channel blockers with mainly vascular effects’ was significantly enriched (FDR = 0.026).

**Figure 4.**
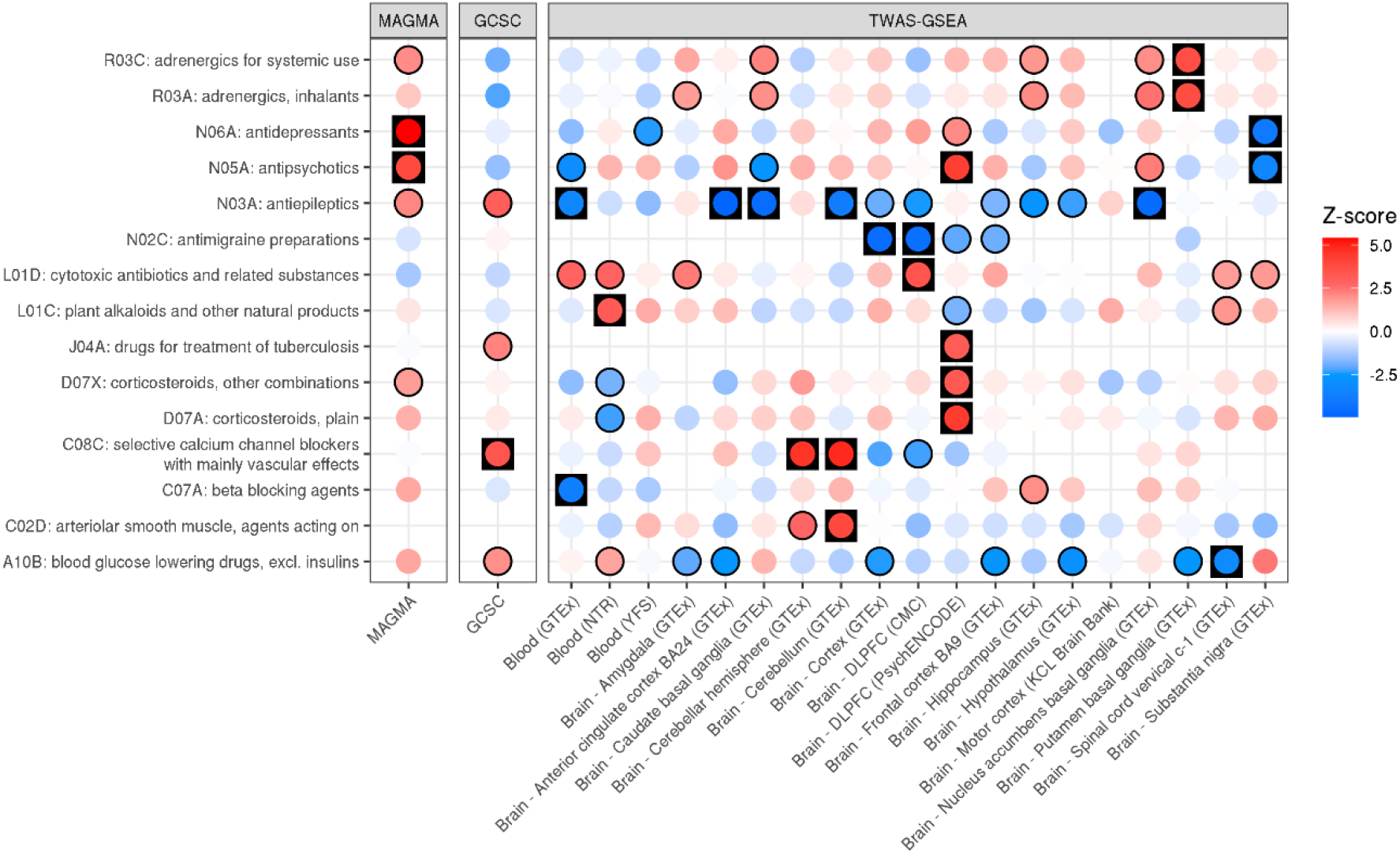
ATC enrichment for ALS based on results of MAGMA, GCSC and TWAS-GSEA. Each point is coloured according to the Z-score of enrichment. For TWAS-GSEA, positive Z-scores (red) indicate the drugs reduces risk of ALS, whereas negative Z-scores (blue) indicate the drugs increases risk of ALS. Results have a black outline if the association was nominally significant, and are in a black square if the association was FDR significant.

Using TWAS-GSEA results there were 22 FDR significant ATC enrichments across all panels, representing 15 unique ATC codes, 4 of which were significant across more than one expression panel. The most enriched ATC group reducing ALS risk was ‘selective calcium channel blockers with mainly vascular effects’, which was FDR significant using two expression panels, consistent with results of GCSC. The most enriched ATC group increasing ALS risk was ‘antiepileptics’, FDR significant across five expression panels. The ‘antipsychotics’ ATC group was significantly enriched using TWAS results from two panels, but the direction of enrichment was opposite: while antipsychotics associated with a decreased ALS risk based on PsychENCODE dorsolateral prefrontal cortex (DLPFC) data, they associated with an increased risk based on GTEx substantia nigra data. Other ATC groups enriched according to TWAS-GSEA are shown in Figure 4.

### Prediction using Polytranscriptomic Scores (PTS)

Using inferred differential gene expression results from the ALS TWAS, we generated PTS in two ALS case-control target samples with observed whole blood expression, stratifying PTS by either brain or blood TWAS gene expression panels, and by evidence of colocalisation (PP4 > 0.8) (Figure 5). We found strong evidence that the ALS PTS can predict ALS case-control status (max. liability *R*^2^ = 4%; *p*-value = 2.1×10^−21^). The ALS PTS also showed evidence of being associated with an increase in likelihood of spinal onset ALS. PTS associations with age of onset and survival time were inconsistent. Partitioning PTS to TWAS associations from either blood or brain expression panels, or by evidence of colocalisation did not substantially affect the predictive utility of the PTS. However, restricting to colocalised associations from blood-based TWAS results consistently reduced the predictive utility of the PTS. PTS association results for each target sample are separately shown in Supplementary Figures 2-5. The number of genes in the PTS available in each target sample varied (Table S7).

**Figure 5.**
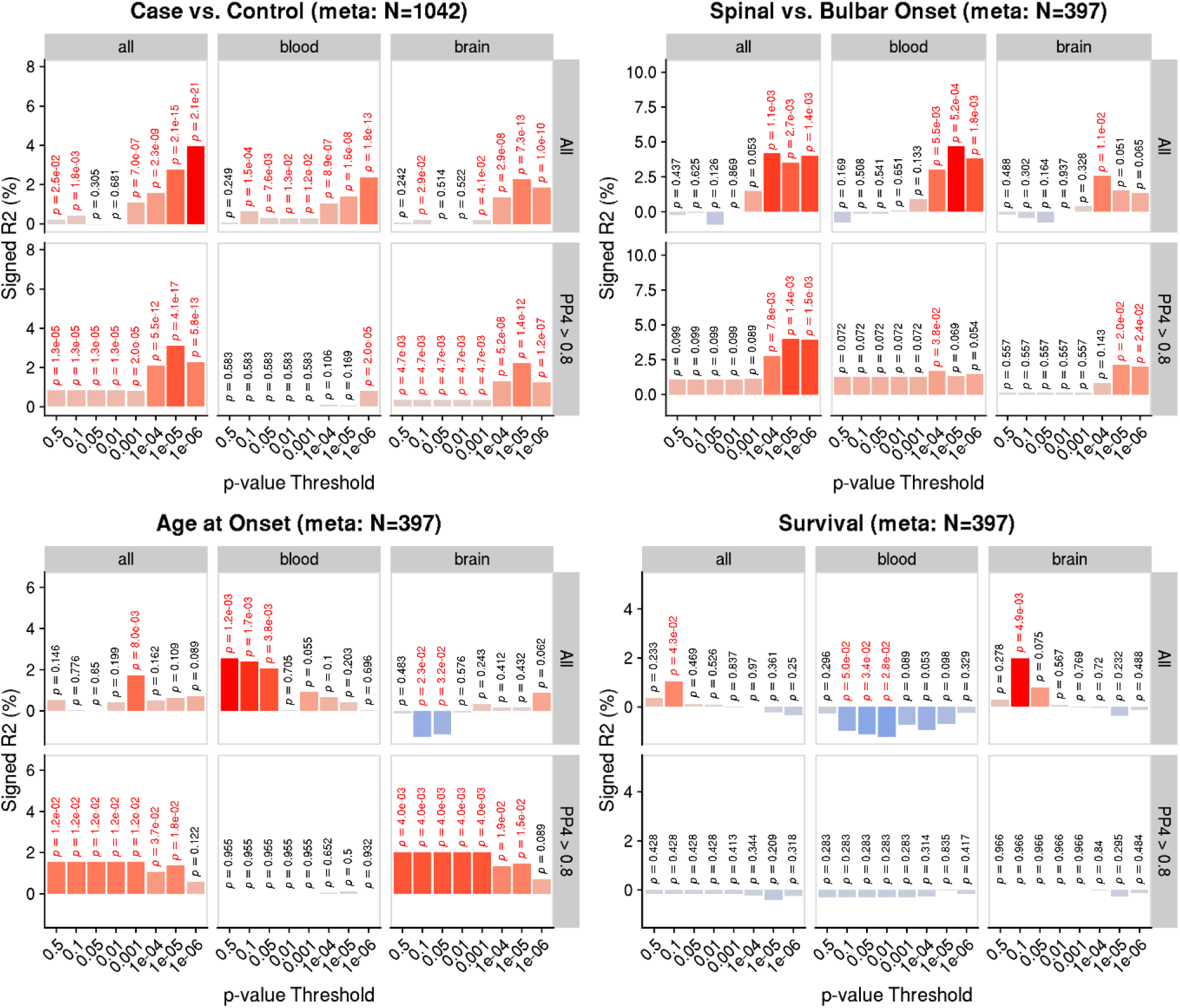
PTS association ALS risk and patient characteristics. Y-axis shows the percentage variance explained signed by the direction of association, on the liability scale for binary outcomes and on observed scale for continuous outcomes. P-values are shown above each bar, with nominally significant (p<0.05) associations highlighted in red. Results are shown when deriving PTS using all TWAS panels, only blood panels and only brain panels. Results are also shown when deriving PTS using only TWAS associations that showed evidence of colocalisation (PP4 > 0.8).

### Comparison of predicted and observed differential expression in ALS

We compared the predicted differential expression from TWAS to observed differential expression in the GPL6947 and GPL10558 ALS case-control blood expression cohorts (Figure 6). Of the 108 high-confidence genes identified using TWAS (FUSION or SMR), 44 genes were statistically significant in the observed differential expression analysis (Figure S6). Of the 36 high-confidence genes from TWAS using blood expression panels specifically, 18 genes had significant observed differential expression, with 8 genes showing a consistent direction of effect (*ATXN3, BTBD1, DHRS11, GGNBP2, MTAP, RNF24, TBK1, ZNHIT3)* and 10 genes showing a discordant direction of effect (*CAMLG, FNBP1, LY6G5C, NCR3, NDST2, PSMB10, PTBP2, SAR1B, SDHA, ZNF142*).

## Discussion

This study has provided novel and mechanistic insights into the genes underlying ALS genetic risk. Furthermore, using these insights, we have identified drugs that could be used to treat ALS and derived an ALS PTS able to predict ALS risk. Collectively these results provide a key advance in our understanding of ALS aetiology, highlight novel research avenues for pharmacological treatment of ALS, and offer a new and non-invasive biomarker of ALS risk.

We used a combination of SNP fine-mapping, TWAS and PWAS analyses to define a set of high-confidence genes associated with ALS risk. By integrating eQTL and pQTL data we have uncovered novel molecular insights regarding ALS, aiding future experimental studies to further characterise the role of specific variants and genes in ALS. For example, we found that ALS risk leads to an increased expression of the *SCFD1* gene in brain tissues, which is consistent with a previous study (Iacoangeli et al., 2021), as well as inferring increased levels of the SCFD1 protein in brain tissues accompanied by a decreased expression of the *SCFD1* gene in the blood. Furthermore, we infer ALS is associated with decreased expression of the *CAMLG* gene, supporting previous findings (Dangond et al., 2004), which we expanded on by showing an increased level of the CAMLG protein in people with ALS. Further validation of this finding is required, but one plausible explanation is that the ALS risk variant increases the post-translational stability of the CAMLG protein, leading to reduced expression of the gene due to a negative feedback loop. Our study also identified many novel ALS genes, such as *TMEM175*. Our study highlights genetic risk for ALS confers a reduced level of the TMEM175 protein in the DLPFC. TMEM175 is a lysosomal ion channel, previously linked to Parkinson’s disease, and a deficiency of TMEM175 has been found to cause decreased mitochondrial respiration, altered lysosomal pH and impaired autophagy (Jinn et al., 2017). Together these findings suggest TMEM175 deficiency may also increase risk of ALS, providing a new insight into the aetiology of ALS.

We then explored several strategies for the identification of drugs that could be repositioned for treating ALS. We leveraged the molecular insights from our TWAS analysis to identify drugs and drug classes that interact with differentially expressed genes in people with ALS, and furthermore indicate whether these drugs might reverse the ALS-risk associated expression profile. We identified five drugs that are enriched for interactions with genes differentially expressed in people with ALS. By using directional information from TWAS and a drug-interaction database, we highlighted that two of the enriched drugs, acarbose and volgibose, will likely exacerbate ALS-risk associated differential expression. These drugs are both α-glucosidase inhibitors used to treat diabetes. This finding suggests opposing pathology of diabetes and ALS, as has been previously indicated using population studies (Dupuis et al., 2008; Kioumourtzoglou et al., 2015; Zhang et al., 2022) and a mendelian randomisation study (Zhang et al., 2022), with less favourable lipid profiles and type II diabetes associated with reduced risk of ALS. However, another study in mice with the c9orf72 mutation indicated the type II diabetes medication metformin might have therapeutic potential for ALS (Zu et al., 2020), indicating ALS risk may be moderated differentially across classes of diabetes medication. Another enriched drug was sertindole, with directional TWAS enrichment suggesting this drug could counteract ALS pathology. Sertindole is an atypical antipsychotic with a strong affinity for dopamine D2, serotonin 5-HT2A, serotonin 5-HT2C, and alpha-1-adrenergic receptors (Lindström & Levander, 2006). It has been found to trigger autophagy in neuronal cells, which can lead to cell death. Further research is needed to understand the potential therapeutic process induced by sertindole.

Beyond enrichment of individual drugs, our analyses provided a broader insight into which classes of drugs might be relevant to ALS. The most strongly enriched drug class predicted to have therapeutic benefits for ALS was selective calcium channel blockers, drugs primarily used to treat hypertension. Several studies show an association between hypertension and ALS, with a recent mendelian randomisation study supporting a causal role between prescription of calcium channel blockers and reduced risk of ALS (Xia et al., 2022), supporting the results of this study and highlighting a novel therapeutic avenue for ALS. In addition, we identified antiepileptic drugs were strongly enriched for interactions with ALS associated genes, however their direction of effect is predicted to exacerbate ALS risk. This finding is novel and should be further investigated to understand the implications for our understanding of ALS aetiology. One antiepileptic drug (ezogabine) has been found to have potential therapeutic effects on ALS (Wainger et al., 2021), so further investigation of which antiepileptics might increase risk for ALS is needed.

To further utilise our novel ALS differential expression results from TWAS, we combined our results with observed blood expression data from people diagnosed with ALS and controls to calculate PTS. We then evaluated whether PTS could predict ALS risk and clinical characteristics. Looking across two independent target expression datasets, the association between ALS PTS and case-control status was highly significant, explaining up to 4% of ALS risk on the liability scale. This suggests blood-based PTS can provide a key advance in our ability to diagnose and predict ALS risk and could be used in combination with other risk factors as a non-invasive clinical biomarker of ALS risk.

Furthermore, the PTS was associated with site of onset, suggesting ALS PTS might help guide the prognosis of people with ALS. This finding also indicates that differential expression inferred by TWAS is concordant with differential expression observed in people with ALS and controls associated with ALS. However, the target sample expression data was collected after diagnosis of ALS and possibly pharmacological treatment, which may confound our PTS association results. Therefore, further research regarding the predictive utility of ALS PTS in other contexts is required.

In addition to evaluating the predictive utility of the PTS, we contrasted the genetically-inferred differential expression from TWAS with *observed* differential expression in the ALS case-control gene expression datasets. Many genes were identified as significantly associated with ALS risk using both approaches, though a large proportion showed discordant directions of effect. Observed differential expression associated with ALS may reflect the gene’s causal role in ALS pathology, but differential expression within people with ALS may also be a consequence of the disease. Given genetically-inferred differential expression is not susceptible to reverse causation, discordance between inferred and observed expression may help distinguish the molecular mechanisms by which these associations occur. A previous study regarding Crohn’s disease demonstrated genes showing discordant genetically-inferred and observed differential expression are more likely to be triggered in response to immune stimuli (Marigorta et al., 2017). A possible example of this from our study is the gene *SAR1B, which* was inferred to be downregulated in those with ALS using TWAS but was observed to upregulated in those with ALS. This could indicate that *SAR1B* is upregulated in those with ALS as a protective response to ALS pathology, and genetic variation in this locus is associated with ALS because it reduces the extent to which *SAR1B* can be upregulated. *SAR1B* has been reported to provide protection against inflammatory processes (Sané et al., 2019), consistent with the notion that *SAR1B* upregulation is a protective response to ALS pathology.

This study should be considered in the context of several limitations. First, in this study we only consider *common* genetic variation associated with ALS. However, the genetic architecture of ALS includes a combination of common and rare genetic variation. Considering the implicated genes and pathways from both sources of ALS risk will provide further insight into possible therapeutic avenues and improve risk prediction. Second, we only consider ALS GWAS summary statistics and functional genomic annotation data based on European ancestry individuals. The expansion of ALS and functional genomic studies in diverse populations will improve the generalisability of the prediction models across populations, as well as strengthen insights into causal mechanisms. Third, although large advances have been made in developing functional genomic datasets, such as eQTL studies, the current samples sizes prohibit the use integration of distal regulatory elements (e.g., trans eQTLs). Furthermore, the eQTL and pQTL datasets used in this study are based on bulk tissue samples, and integration of single cell-based panels in the future will enable further mechanistic understanding of ALS (Yao et al., 2020).

In conclusion, this study provides novel mechanistic insights into the genes associated with ALS, drug enrichment analysis has highlighted several therapeutic research avenues, and our findings indicate PTS may be a powerful predictor of ALS risk.

## Data Availability

All data produced in the present study are available upon reasonable request to the authors

## Acknowledgements

The authors acknowledge use of the research computing facility at King’s College London, Rosalind (https://rosalind.kcl.ac.uk), which is delivered in partnership with the NIHR Maudsley BRC, and part-funded by capital equipment grants from the Maudsley Charity (award 980) and Guy’s & St. Thomas’ Charity (TR130505). The views expressed are those of the authors and not necessarily those of the NHS, the NIHR or the Department of Health and Social Care.

## Supplementary Information

### 1. GWAS quality control

GWAS summary statistics underwent quality control to extract variants present in the 1000 Genomes (1KG) phase 3 reference sample (1000 Genomes Project Consortium, 2015), remove ambiguous variants, remove variants with missing data, flip variants to match the reference, retain variants with a minor allele frequency (MAF) > 0.01 in the European subset of 1KG Phase 3, retain variants with a MAF > 0.01 in the GWAS sample, remove variants with a discordant MAF (> 0.2) between the reference and GWAS sample, remove variants with association *p*-values >1 or ≤ 0, remove duplicate variants, and remove variants with sample size > 3SD from the median sample size.

### 2. Fine-mapping

Independent genome-wide significant loci were defined using LD-based clumping (*r*^2^ < 0.1, 500kb window), applying SuSiE to all variants within 500kb of each lead variant.

Fine-mapping is particularly sensitive to LD mismatch between GWAS summary statistics and the LD reference. Given we did not have estimates of LD from the original samples in the ALS GWAS, we set the number of causal signals within each locus to 1 (i.e., L = 1), as when L = 1 fine-mapping does not consider LD estimates at all and is therefore more robust. The limitation of the L = 1 assumption is that fine-mapping will be less powerful in loci where multiple causal signals are present.

As a sensitivity analysis, we ran fine-mapping using the default L = 10 parameter, allowing up to 10 independent causal signals within each locus, using the European ancestry subset of the 1KG reference to calculate LD.

### 3. KCL Brain Bank

The KCL Brain Bank dataset previously underwent quality control and standard eQTL analysis (Iacoangeli et al., 2021; Jones et al., 2021). We used a broader version of the dataset to previous publications without age and sex matching between individuals with and without ALS diagnosis as we are not trying to identify associations with ALS. Using GenoPredPipe (https://github.com/opain/GenoPred/tree/master/GenoPredPipe) and the 1KG Phase 3 reference, we identified individuals in KCL Brain Bank of European ancestry and calculated 10 genetic principal components. The final dataset consisted of 153 individuals, including 103 individuals diagnosed with ALS and 50 controls. The expression data was controlled for ALS status, gender, age, post-mortem delay, RIN (RNA integrity), surrogate variables and genetic principal components. We used only HapMap3 variants when generating TWAS weights to improve overlap with external datasets. The weights were created using FUSION software, implemented using a publicly available pipeline (https://github.com/opain/Calculating-FUSION-TWAS-weights-pipeline).

Post-mortem tissue samples from King’s College London were collected under the ethical approval of the MRC London Neurodegenerative Diseases Brain Bank and under the regulations of the Human Tissue Act UK 2014. All post-mortem tissue was donated to the MRC London Neurodegenerative Diseases Brain Bank under standard ethical and Human Tissue Act procedures, with informed consent provided by the next of kin. Data generated from this material were anonymized and analysed on a high-performance computing cloud (https://www.maudsleybrc.nihr.ac.uk/facilities/rosalind/)with data protection protocols in accordance with Department of Health Policy (UK) and the security standards set by the National Data Guardian. Ethical approval to process and analyse post-mortem samples stored at King’s College London was provided by a local ethics committee at the Institute of Psychiatry, Psychology & Neuroscience, King’s College London, and the MRC London Neurodegenerative Diseases Brain Bank.

### 4. FUSION/SMR

FUSION software implements an approach often referred to as TWAS, which infers differential expression/protein levels associated with the GWAS phenotype using multi-variant models predicting expression or protein levels (Gusev et al., 2016). These multi-variant models are not available for all eQTL datasets, partly because deriving these multi-variant models currently requires individual-level gene expression and genotype data.

SMR also infers differential expression/protein levels associated with the GWAS phenotype (Zhu et al., 2016), but aims to provide evidence for causal role of a given SNP on a trait mediated through gene expression. SMR only considers the genetic variant most strongly predicting expression or protein levels, thereby explaining less variance in expression/protein levels than FUSION multi-SNP models for genes which have secondary eQTL/pQTL effects. A current advantage of SMR over FUSION is that it can be applied using only eQTL summary statistics which are more widely available.

Both FUSION and SMR include an analysis to determine whether the overlapping genetic association for the phenotype and the gene expression is driven by the same causal variant (pleiotropy) or whether different causal variants that are in LD are driving the associations (linkage). FUSION uses the coloc package to perform Bayesian colocalisation (Giambartolomei et al., 2014). SMR uses the frequentist HEIDI test.

### 5. Drug enrichment methods

MAGMA gene-set enrichment analysis is based on MAGMA estimated gene associations and binary drug-gene interaction data (de Leeuw et al., 2015). This approach does not consider the direction of ALS-gene or drug-gene associations, so it identifies drugs that interact with genes enriched for association with ALS but does not indicate whether enriched drugs will decrease risk of ALS. MAGMA gene set enrichment analysis estimates the non-independence of gene associations by using an LD-based correlation matrix and a generalised least square model.

Gene co-regulation score (GCSC) regression is a method that leverages gene co-regulation to test for and enrichment of TWAS associations within gene-sets or associated with gene-properties (Siewert-Rocks et al., 2022). Like MAGMA, GCSC does not consider the direction of effect between the gene and the phenotype. GCSC was run using default settings and the publicly available co-regulation matrices, based on GTEx v7 expression (https://github.com/ksiewert/GCSC). We restricted the analysis to GTEx brain tissues and GTEx whole blood, consistent with gene expression panels included in our TWAS analysis. As currently required for use of the GCSC coregulation matrices, we performed a TWAS using the GTEx v7 expression panels as input for the GCSC analysis.

TWAS-based gene-set enrichment analysis (TWAS-GSEA) is based on TWAS estimated gene association and directional drug-gene interaction data (Pain et al., 2019). This approach does consider the direction of ALS-gene and drug-gene associations, so enriched drugs using this method are suggested to induce expression changes reducing risk of ALS. TWAS-GSEA estimates the non-independence of gene associations using a predicted expression correlation matrix and a linear mixed model, using the lme4qtl R package (Ziyatdinov et al., 2018).

These three approaches have different advantages and limitations and are therefore complimentary. MAGMA gene associations have no clear mechanistic link to the phenotype and the enrichment analysis does not consider direction of effect. However, MAGMA will generally test for enrichment across more genes that TWAS-based enrichment as it considers ALS-associated genes acting via any mechanism (not only differential expression) and is not dependent on external eQTL datasets which are often limited in sample size thereby reducing coverage of the genome. GCSC has the advantage of pooling expression associations across expression panels, thereby improving coverage and statistical power to detect enrichment. TWAS-GSEA does not pool information across expression panels but does allow for the direction of effect in the TWAS to be considered, and is thereby able to highlight drugs that consistently reduce risk-associated differential expression.

### 6. Gene discovery results

SNP-based fine-mapping results assuming a single causal signal (L = 1) are summarised in Table S1. LD-based clumping identified 16 independent genome-wide significant associations, which were carried forward for fine-mapping analysis centred on the lead variant +/- 500kb. Within three loci a single variant was present in the 95% credible set. Within four loci the 95% credible set was contained within a given gene, with one 95% credible set within two overlapping genes. SNP-based fine-mapping results allowing for up to 10 causal signals (L = 10) are shown in Table S2, indicating the presence of multiple causal signals underlying genome-wide significant locus on chromosome 9, with 95% credible sets contained within the *C9orf72* and *MOB3B* genes.

TWAS using FUSION identified 197 FDR-significant and colocalised associations for ALS, including 101 unique genes (Table S3). TWAS using SMR identified 44 FDR significant associations passing the HEIDI test (indicating colocalisation), including 29 unique genes (Table S4). Across FUSION and SMR, 108 unique genes were identified as significant and colocalised, of which 22 were found by both TWAS and SMR.

PWAS using FUSION with ROSMAP and Banner pQTL data identified 8 FDR-significant and colocalised associations, including 6 unique genes, and thereby 2 were significant and colocalised according to both ROSMAP and Banner (Table S5). SMR using ROSMAP pQTL data identified 4 significant and colocalised associations, of which 3 were in common with FUSION PWAS results (Table S6). Of the 7 unique genes implicated using PWAS analysis, 3 were in common with TWAS results.

Across SNP fine-mapping, TWAS and PWAS analyses, 117 unique genes were identified as high-confidence associations.

## Supplementary Figures

**Figure S1a.**
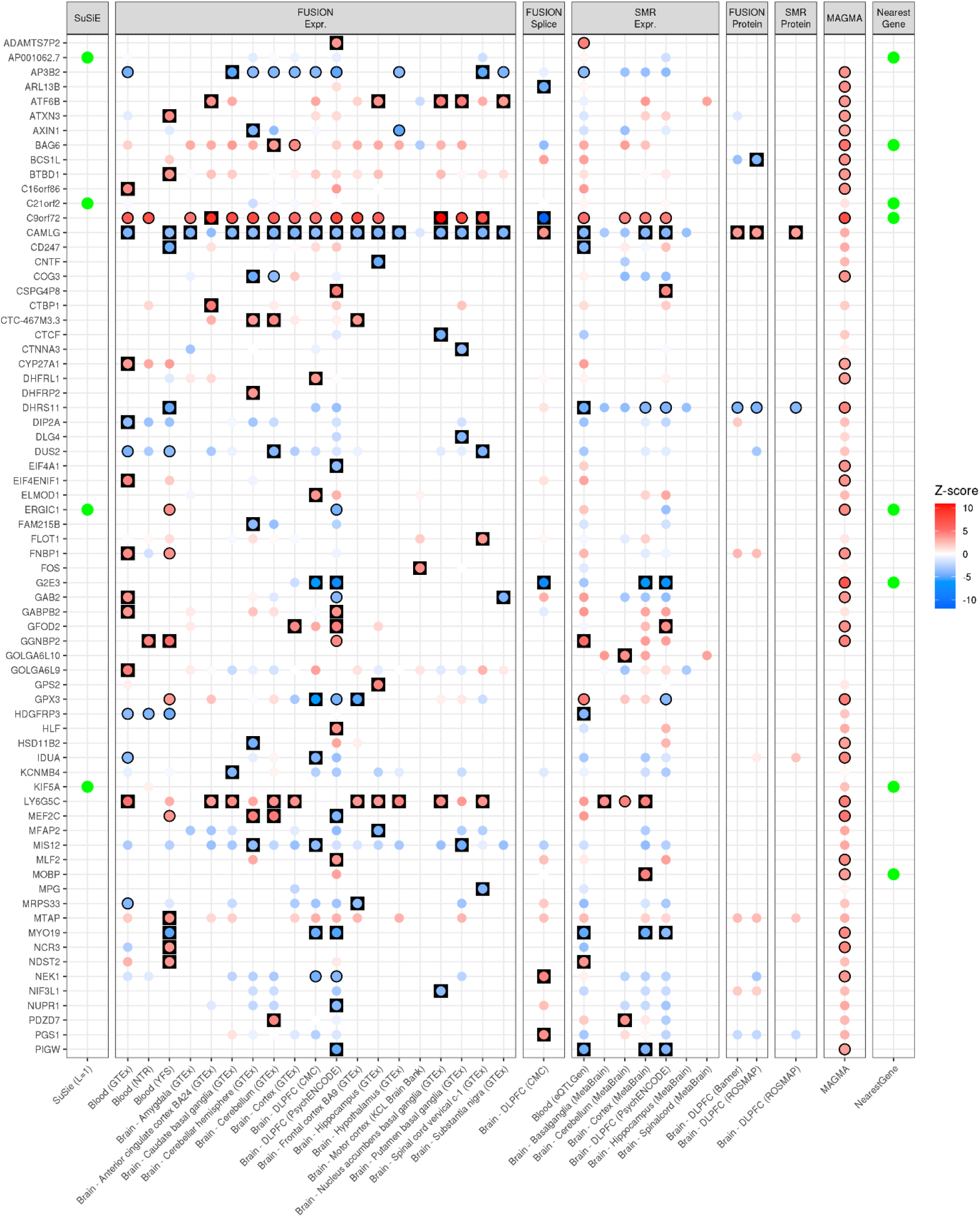
High-confidence gene list for ALS defined using results of SNP fine-mapping, TWAS and PWAS. MAGMA and NearestGene results included for comparison. Results are separated by the method and external data used. Genes containing the 95% credible set from SNP fine-mapping (L = 1) are indicated using a green box. FUSION and SMR results are shown for all panels, with each boxed coloured according to the Z-score of association. Red indicates an increased expression/protein level in people diagnosed with ALS, and blue indicates decreased expression/protein level in people diagnosed with ALS. FUSION and SMR results have a black outline if the association was FDR significant, and are in a black square if the association was FDR significant and showed evidence of colocalisation. MAGMA associations are also shaded according to Z-score, although MAGMA cannot infer the direction of the effect, with FDR significant genes outlined in black. Genes nearest to lead variants within genome-wide significant loci are indicated using a green point.

**Figure S1b.**
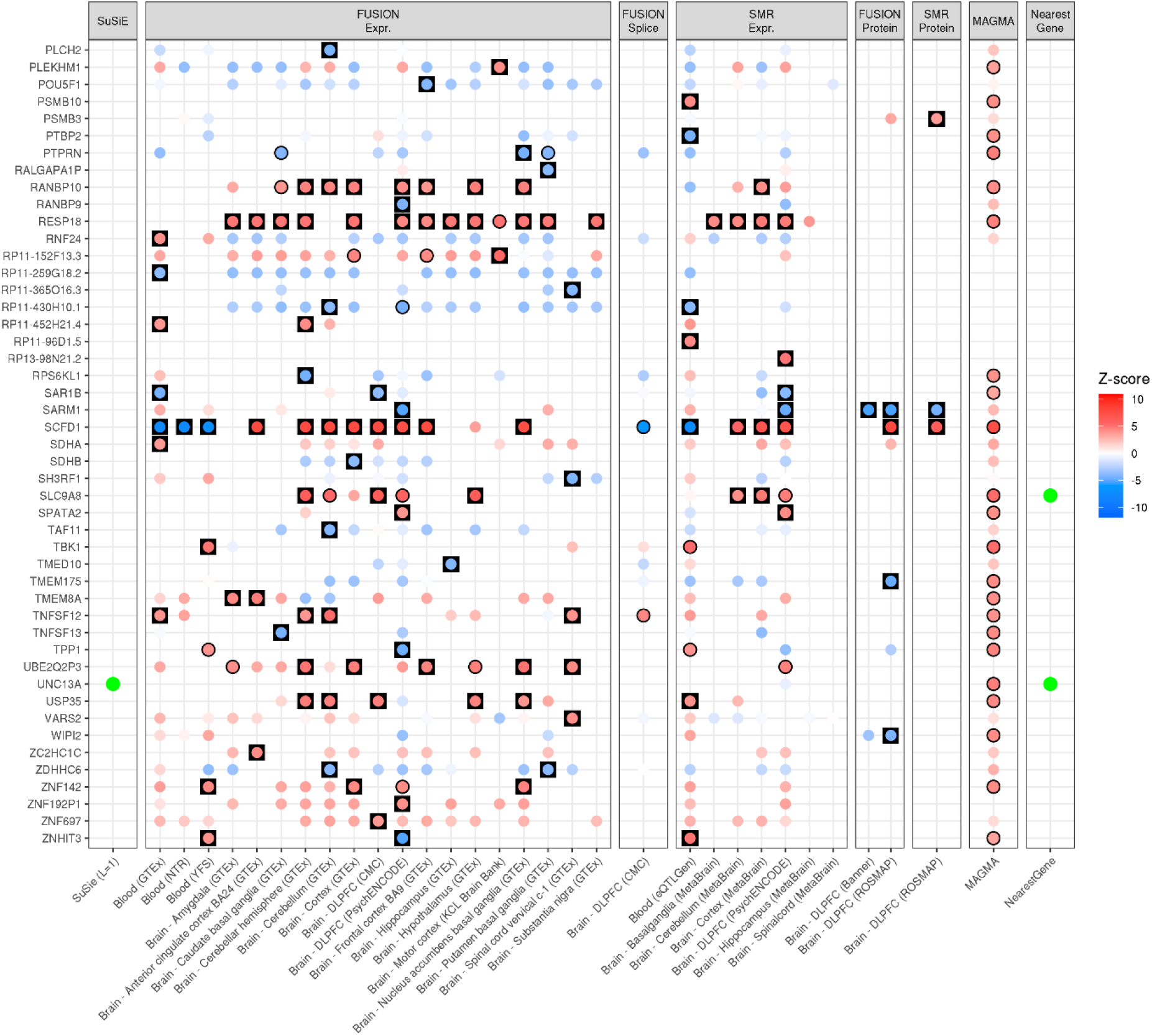
High-confidence gene list for ALS defined using results of SNP fine-mapping, TWAS and PWAS. MAGMA and NearestGene results included for comparison. Results are separated by the method and external data used. Genes containing the 95% credible set from SNP fine-mapping (L = 1) are indicated using a green box. FUSION and SMR results are shown for all panels, with each boxed coloured according to the Z-score of association. Red indicates an increased expression/protein level in people diagnosed with ALS, and blue indicates decreased expression/protein level in people diagnosed with ALS. FUSION and SMR results have a black outline if the association was FDR significant, and are in a black square if the association was FDR significant and showed evidence of colocalisation. MAGMA associations are also shaded according to Z-score, although MAGMA cannot infer the direction of the effect, with FDR significant genes outlined in black. Genes nearest to lead variants within genome-wide significant loci are indicated using a green point.

**Figure S2.**
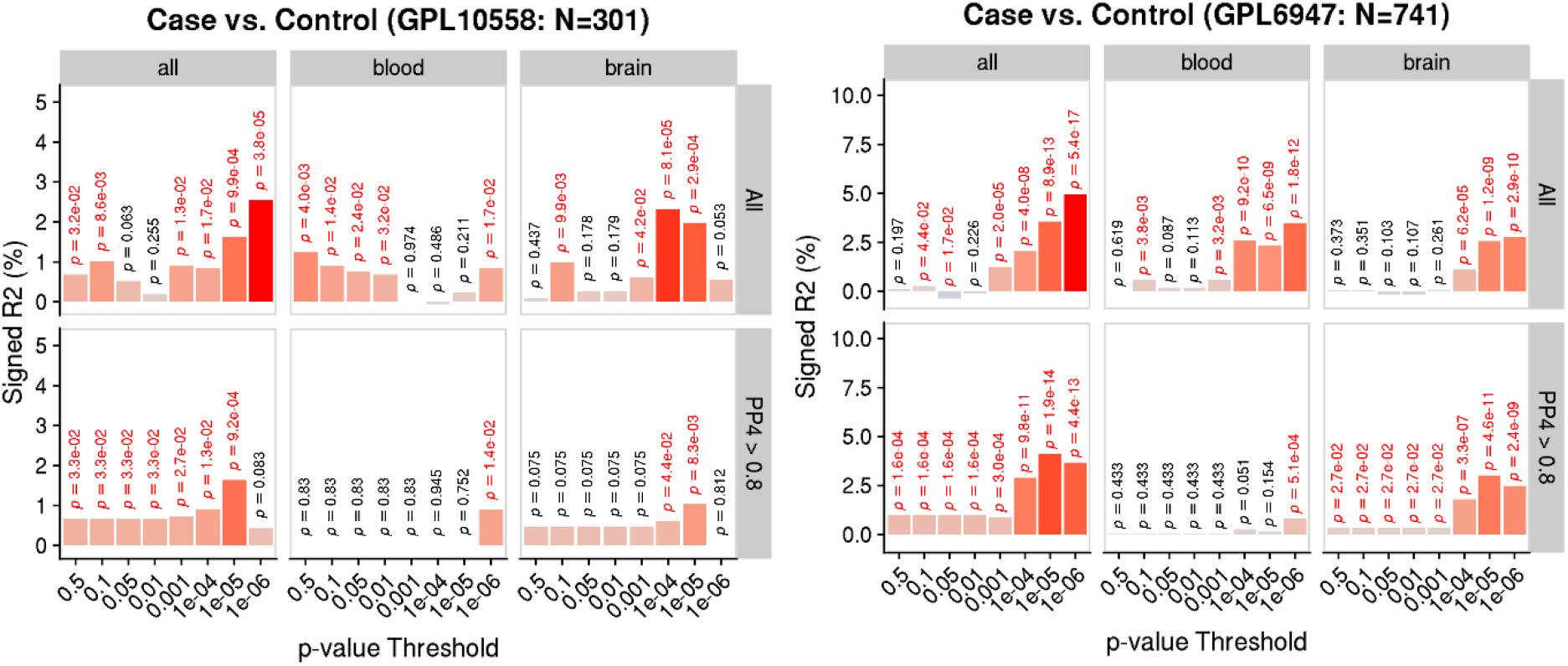
PTS association with ALS cases vs. controls split by target expression platform. Y-axis shows the variance explained on the liability scale assuming a prevalence of 1/300, signed by the direction of association. P-values are shown above each bar, with nominally significant associations highlighted in red. Results are shown when deriving PTS using all TWAS panels, only blood panels and only brain panels. Results are also shown when deriving PTS using only TWAS associations that showed evidence of colocalisation (PP4 > 0.8).

**Figure S3.**
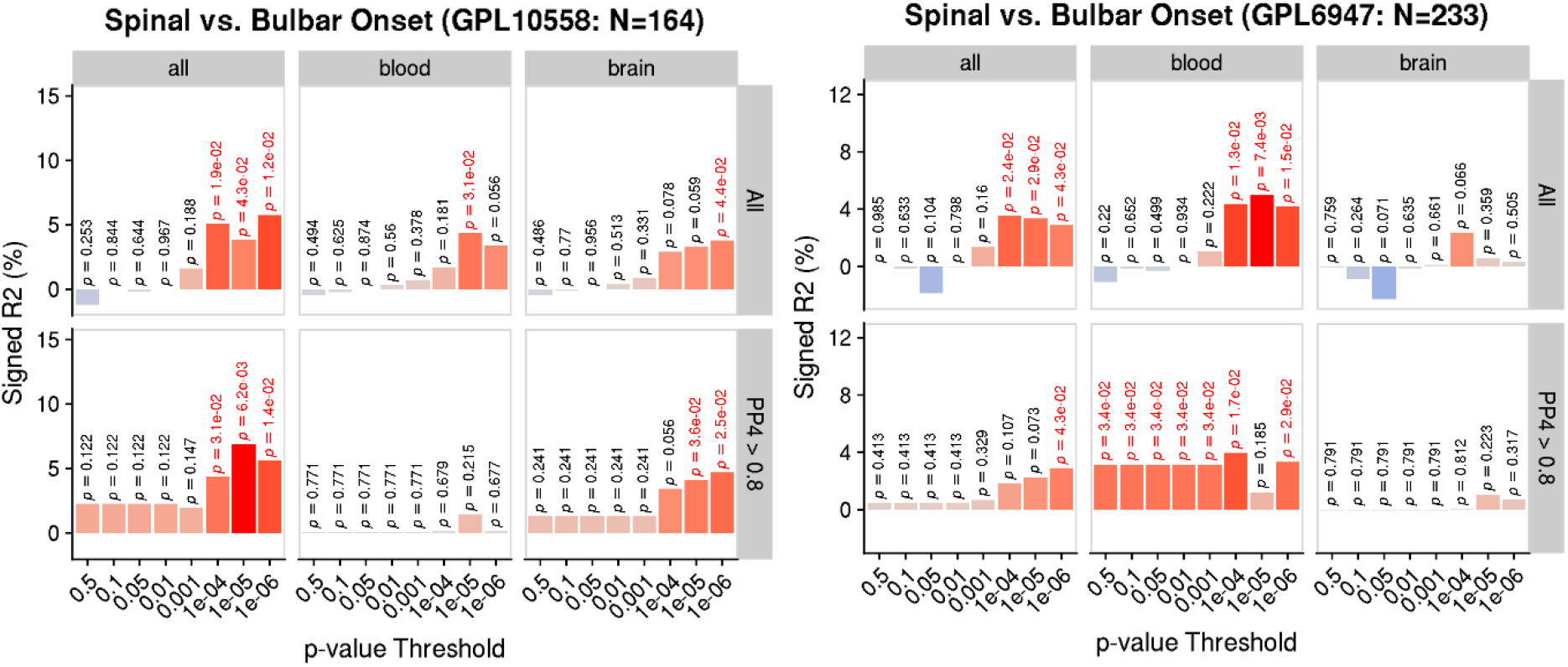
PTS association with site of onset in people with ALS split by target expression platform. Y-axis shows the variance explained on the liability scale assuming a prevalence of 50%, signed by the direction of association. P-values are shown above each bar, with nominally significant associations highlighted in red. Results are shown when deriving PTS using all TWAS panels, only blood panels and only brain panels. Results are also shown when deriving PTS using only TWAS associations that showed evidence of colocalisation (PP4 > 0.8).

**Figure S4.**
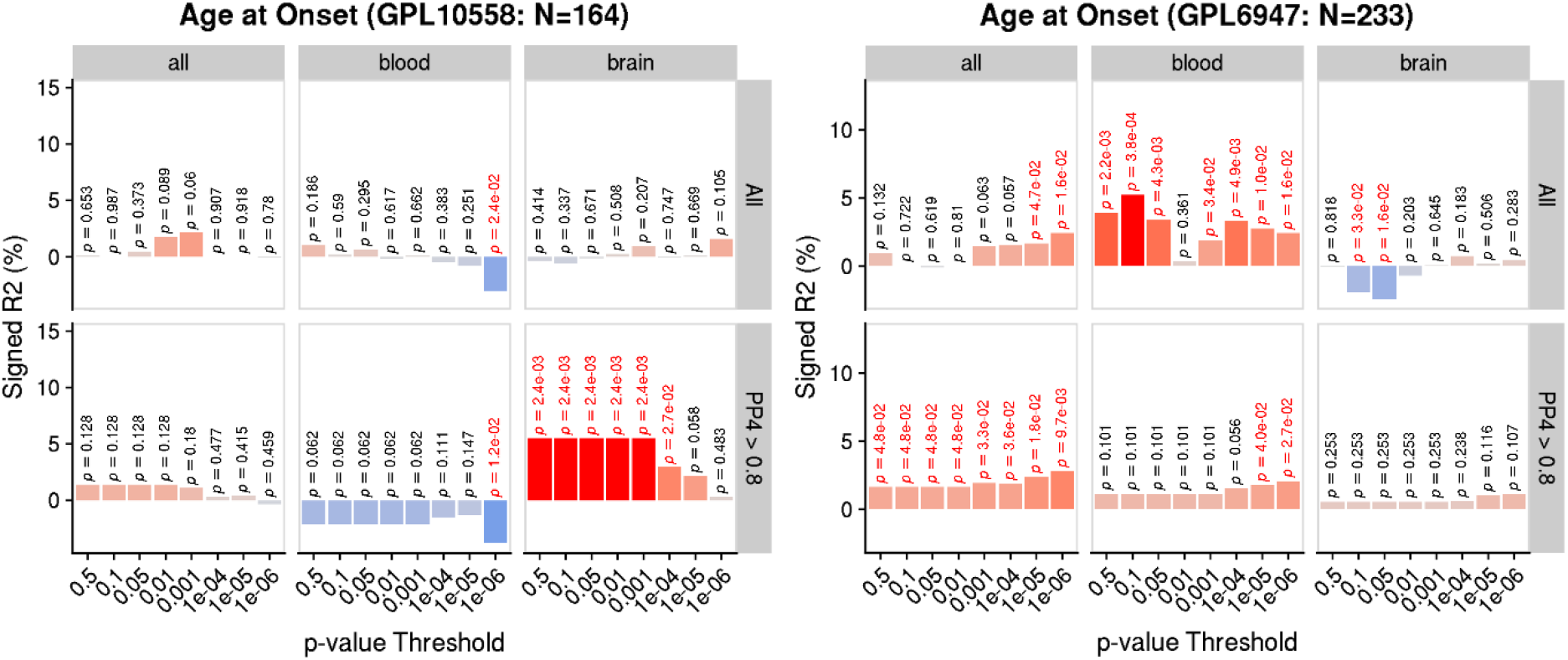
PTS association with age of onset in people with ALS split by target expression platform. Y-axis shows the variance explained on the observed scale, signed by the direction of association. P-values are shown above each bar, with nominally significant associations highlighted in red. Results are shown when deriving PTS using all TWAS panels, only blood panels and only brain panels. Results are also shown when deriving PTS using only TWAS associations that showed evidence of colocalisation (PP4 > 0.8).

**Figure S5.**
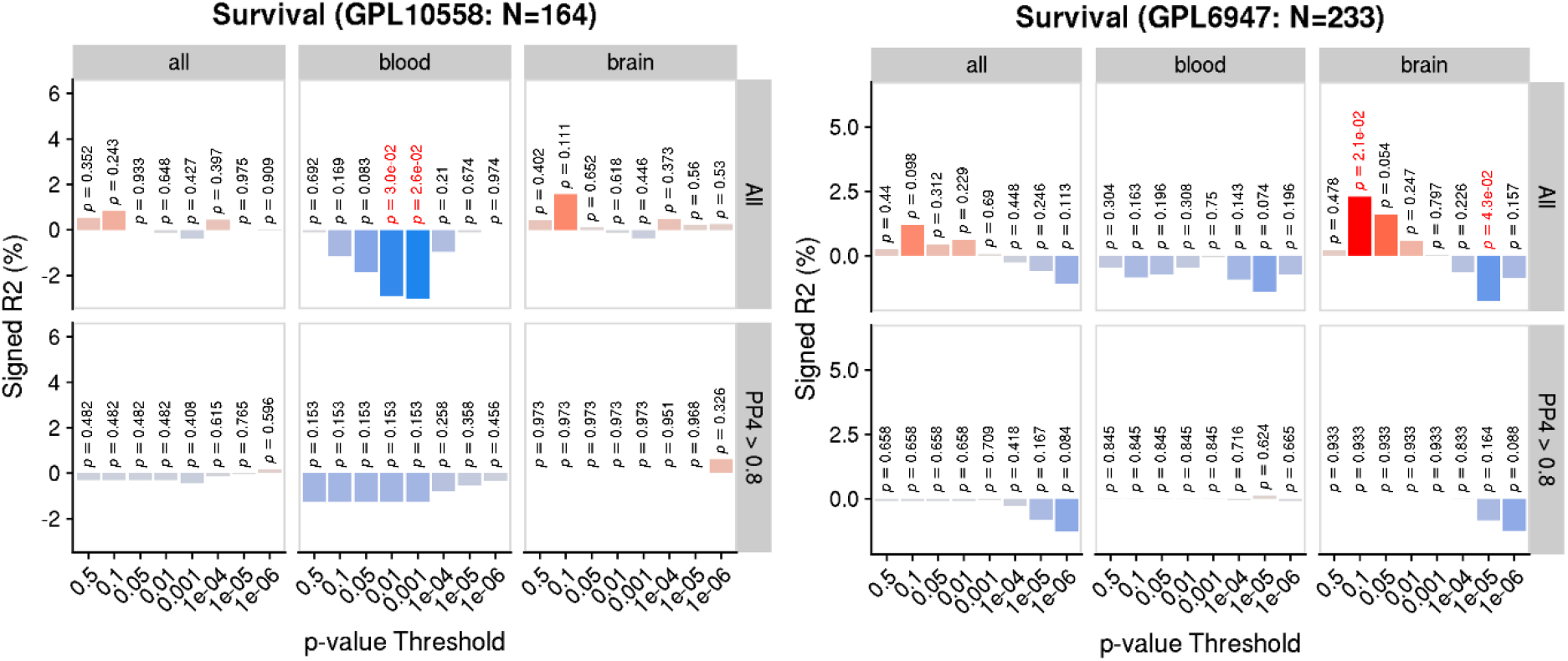
PTS association with site of onset in people with ALS split by target expression platform. Y-axis shows the variance explained on the oserved scale, signed by the direction of association. P-values are shown above each bar, with nominally significant associations highlighted in red. Results are shown when deriving PTS using all TWAS panels, only blood panels and only brain panels. Results are also shown when deriving PTS using only TWAS associations that showed evidence of colocalisation (PP4 > 0.8).

**Figure S6.**
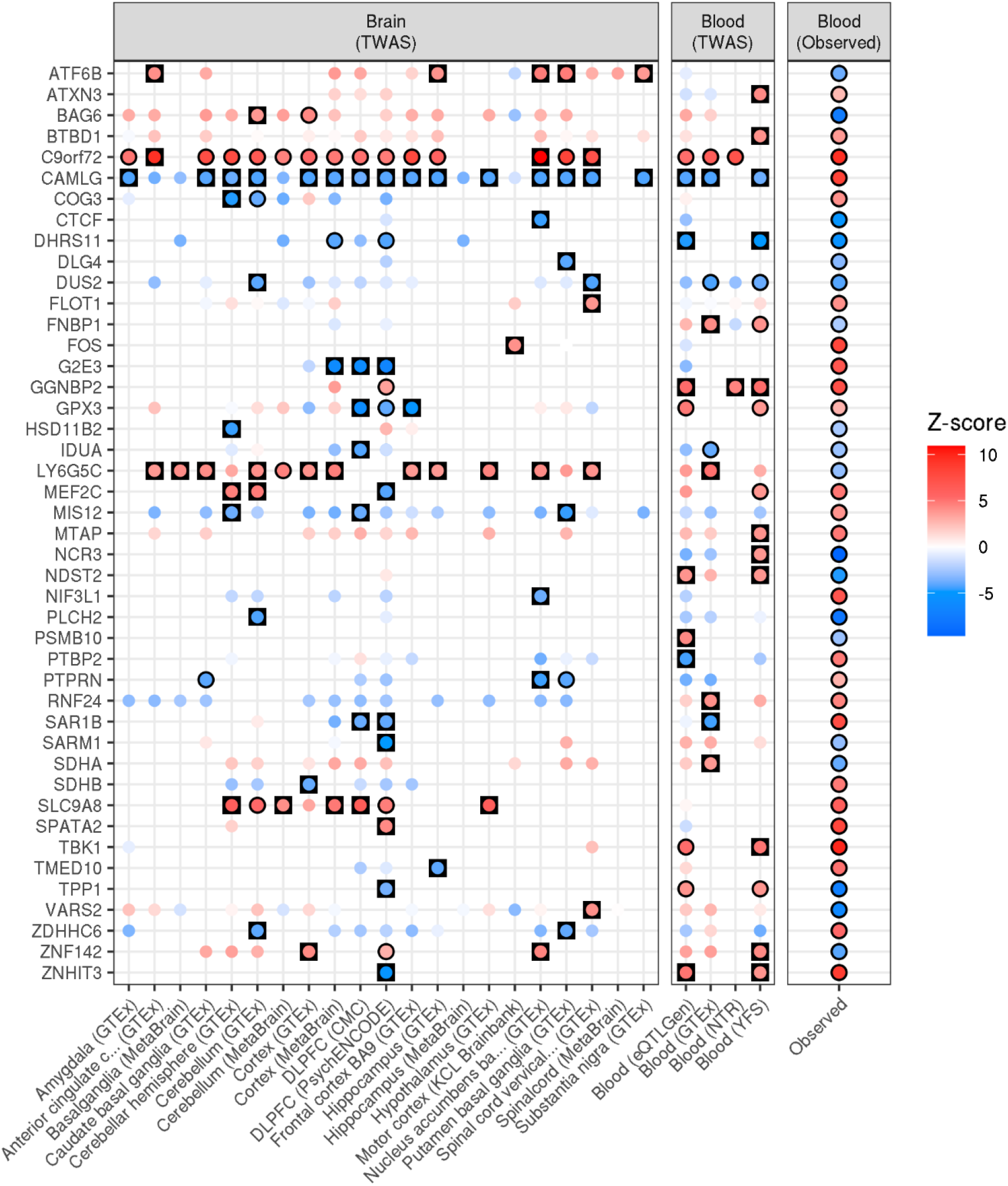
Comparison of observed and predicted (TWAS) differential expression associated with ALS risk. Showing only genes that were identified as high-confidence using TWAS (FUSION or SMR) and FDR significant observed differential expression. Results have a black outline if the association was FDR significant. TWAS results are highlighted in a black square if the association was FDR significant and showed evidence of colocalisation.

